# SARS-CoV-2 RNA detected in blood samples from patients with COVID-19 is not associated with infectious virus

**DOI:** 10.1101/2020.05.21.20105486

**Authors:** Monique I Andersson, Carolina V Arancibia-Cárcamo, Kathryn Auckland, J Kenneth Baillie, Eleanor Barnes, Tom Beneke, Sagida Bibi, Miles Carroll, Derrick Crook, Kate Dingle, Christina Dold, Louise O Downs, Laura Dunn, David W Eyre, Javier Gilbert Jaramillo, Heli Harvala, Sarah Hoosdally, Samreen Ijaz, Tim James, William James, Katie Jeffery, Anita Justice, Paul Klenerman, Julian Knight, Michael Knight, Xu Liu, Sheila F Lumley, Philippa C Matthews, Anna L McNaughton, Alexander J Mentzer, Juthathip Mongkolsapaya, Sarah Oakley, Marta S Oliveira, Timothy Peto, Rutger J Ploeg, Jeremy Ratcliff, David J Roberts, Justine Rudkin, Rebecca A Russell, Gavin Screaton, Malcolm G Semple, Donal Skelly, Peter Simmonds, Nicole Stoesser, Lance Turtle, Sue Wareing, Maria Zambon

## Abstract

**Background:** Laboratory diagnosis of SARS-CoV-2 infection (the cause of COVID-19) uses PCR to detect viral RNA (vRNA) in respiratory samples. SARS-CoV-2 RNA has also been detected in other sample types, but there is limited understanding of the clinical or laboratory significance of its detection in blood.

**Methods:** We undertook a systematic literature review to assimilate the evidence for the frequency of vRNA in blood, and to identify associated clinical characteristics. We performed RT-PCR in serum samples from a UK clinical cohort of acute and convalescent COVID-19 cases (n=212), together with convalescent plasma samples collected by NHS Blood and Transplant (NHSBT) (n=111 additional samples). To determine whether PCR-positive blood samples could pose an infection risk, we attempted virus isolation from a subset of RNA-positive samples.

**Results:** We identified 28 relevant studies, reporting SARS-CoV-2 RNA in 0-76% of blood samples; pooled estimate 10% (95%CI 5-18%). Among serum samples from our clinical cohort, 27/212 (12.7%) had SARS-CoV-2 RNA detected by RT-PCR. RNA detection occurred in samples up to day 20 post symptom onset, and was associated with more severe disease (multivariable odds ratio 7.5). Across all samples collected ≥28 days post symptom onset, 0/143 (0%, 95%CI 0.0-2.5%) had vRNA detected. Among our PCR-positive samples, cycle threshold (ct) values were high (range 33.5-44.8), suggesting low vRNA copy numbers. PCR-positive sera inoculated into cell culture did not produce any cytopathic effect or yield an increase in detectable SARS-CoV-2 RNA.

**Conclusions:** vRNA was detectable at low viral loads in a minority of serum samples collected in acute infection, but was not associated with infectious SARS-CoV-2 (within the limitations of the assays used). This work helps to inform biosafety precautions for handling blood products from patients with current or previous COVID-19.

## BACKGROUND

Since January 2020, the SARS-CoV-2 virus has caused a global pandemic of COVID-19, challenging hospitals and laboratory services worldwide [1]. Diagnosis of infection has largely been based on RT-PCR amplification of viral nucleic acid from the upper respiratory tract (nose/throat) swabs [2]. However, detection of viral RNA (vRNA) has also been reported in blood, serum and plasma from clinical small case series (e.g. [3,4]). The frequency and quantification of SARS-CoV-2 RNA in blood fractions, and the significance of blood as a transmission route remains unknown.

Understanding the clinical contexts within which SARS-CoV-2 RNA can be detected in blood is important to determine the extent to which PCR-positive blood, plasma or serum could have impact as a clinically useful biomarker of disease severity or prognosis. Furthermore, there is an urgent need to consider whether the detection of viral RNA in blood samples reflects the presence of infectious virus, as this has important safety implications for clinicians and laboratory personnel engaged in both routine laboratory testing, as well as COVID-19-specific pipelines such as serology [5,6].

Different organisations have made varying recommendations for the laboratory handling of samples from patients with suspected or confirmed SARS-CoV-2, but these have had to be developed quickly in the face of little experience or data, and rely on the presence of viral RNA in samples as an imperfect surrogate for live virus. Laboratory protocols seeking to reduce the bioburden of SARS-CoV-2 in clinical samples suggest either chemical inactivation (e.g. with sodium-dodecyl-sulfate, Triton-X100, and/or guanidinium thiocyanate-lysis buffers), alone or in combination with heating protocols that vary from 30°C up to as high as 92°C for 15 minutes [7]. These approaches add processing time, may require additional laboratory reagents, and are also potentially associated with a loss of sensitivity in any downstream analysis, particularly pertinent for serological assays. Previous reports suggest that heat inactivation may be particularly detrimental to the sensitivity of antibody detection [8].

An alternative to chemical or heat inactivation is to undertake all sample handling in a biosafety (containment) level 3 (BSL3) facility, but this is expensive, requires specialist staff training, substantially reduces the number of samples that can be processed, and is completely inaccessible in many settings. There is a lack of consensus about appropriate biosafety precautions, and escalate to BSL3 may be based on concerns about risks associated with viraemic samples even when the risk of aerosol generation is low, and there are no data to suggest a risk of blood-borne transmission to laboratory staff.

Here we assimilate the peer-reviewed literature describing the presence of SARS CoV-2 RNA in human blood, with the aim of providing a pooled dataset to provide improved insights into the causes and correlates of RNA-aemia. We then present our own investigation of the frequency and determinants of vRNA detection in blood using 424 samples collected from acutely infected and convalescent patients infected with SARS-CoV-2. We attempted *in vitro* isolation of the virus from viraemic samples in order to determine whether RNA detection is a marker of infectious virus. Together, these data may help to determine the significance of viral RNA in blood, and can contribute to the development of consistent and evidence-based laboratory protocols.

## METHODS

### Terminology and definitions

- **Blood:** we have used the term blood to refer to whole blood, serum or plasma when there is not a clear distinction in existing pre-published data, although we recognise that there may be differences in the sensitivity of viral detection between whole blood and blood fractions.
- **Serum:** in the work undertaken here, we refer specifically to serum, as this blood fraction was consistently used across our experiments.
- **RNA-aemia:** we have used this term to describe the presence of viral RNA, above the technical limits of detection of RT-PCR assays, in blood, serum or plasma. The alternative term, ‘viraemia’, suggests the presence of whole virus in blood. Since we have not demonstrated the presence of replication-competent (infectious) SARS-CoV-2 in the blood compartment, we have elected to use the more conservative description of RNA-aemia (which may or not indicate viraemia).

### Systematic literature review

We searched PubMed, Web of Science, MedRxiv and Google between 7th-11th May 2020, using the search terms (“SARS-CoV-2” OR “COVID” OR “2019-nCoV” OR “COVID-19” OR “2019 NCOV” or “SARS COV 2” or “2019NCOV” or “2019-nCoV” or “2019 novel coronavirus”) AND (“qPCR” OR “RT-PCR” OR “PCR” OR “VIRAL LOAD” OR “RNAaemia” OR “RNAemia” OR “viraemia” OR “viremia” OR “RNA-aemia” OR “RNA-emia”) AND (“BLOOD” OR “PLASMA” OR “SERUM”). We excluded animal studies. We did not make exclusions on the basis of language, but two papers not in English were ruled out because they did not contain details of vRNA detection that we required. Each study was reviewed by at least two independent reviewers. A PRISMA flow chart is presented, showing identification of 28 relevant studies (Figure 1; Table S1). We collected information on the prevalence of vRNA detection in blood, serum or plasma, noting whether this attribute was correlated with clinical or laboratory phenotypes of disease, and recording Ct values when these were reported. Data were collated in Microsoft Excel. To undertake a meta-analysis, we removed one study each of uninfected (healthy) donors and convalescent individuals, and four studies with <5 participants, taking the final number of studies analysed to 22 (Figure 2).

**Figure 1.**
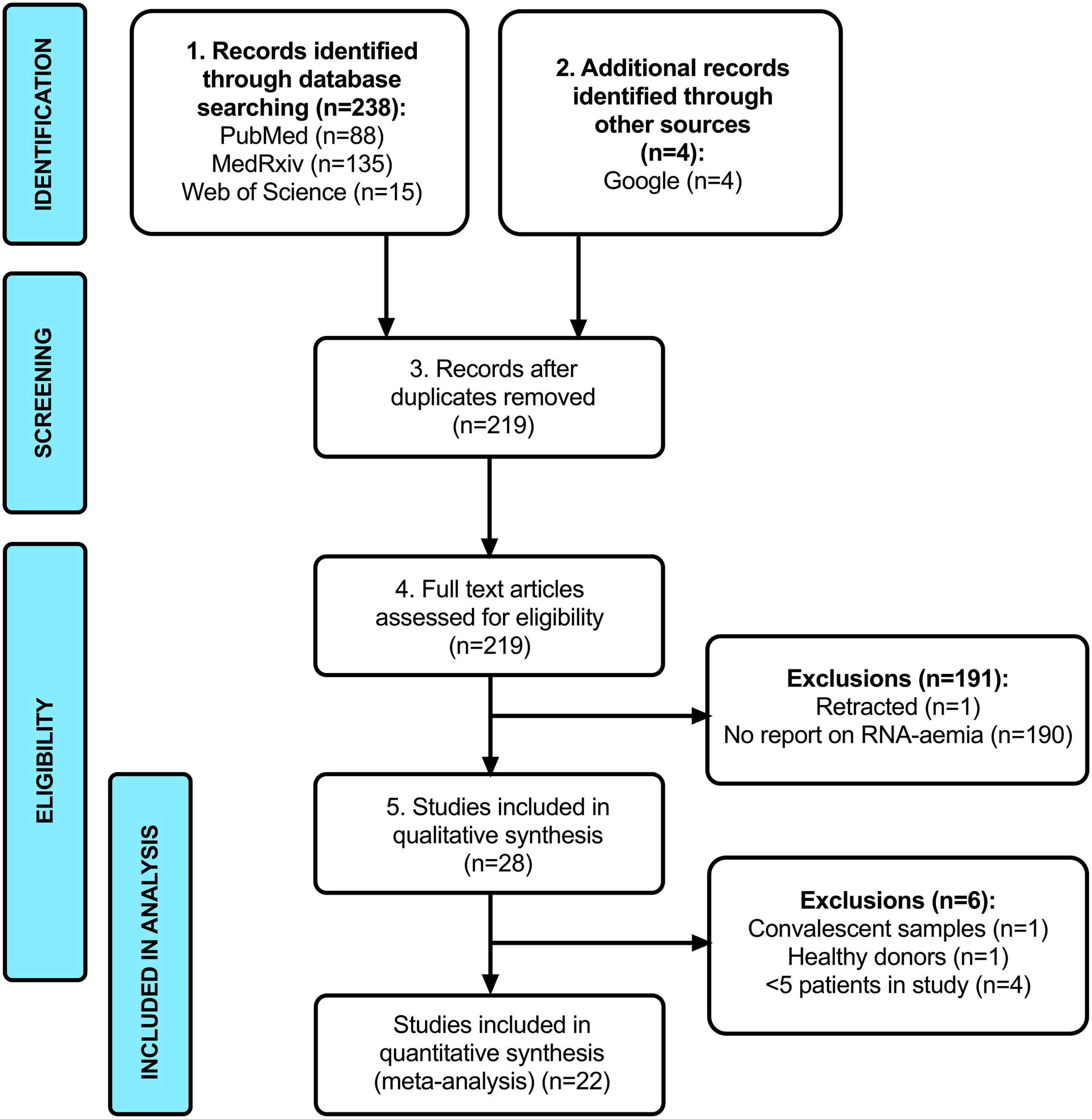
PRISMA flow diagram showing number of abstracts identified through a systematic literature review, rejections (with reasons), and final number of studies included in the analysis.

**Figure 2.**
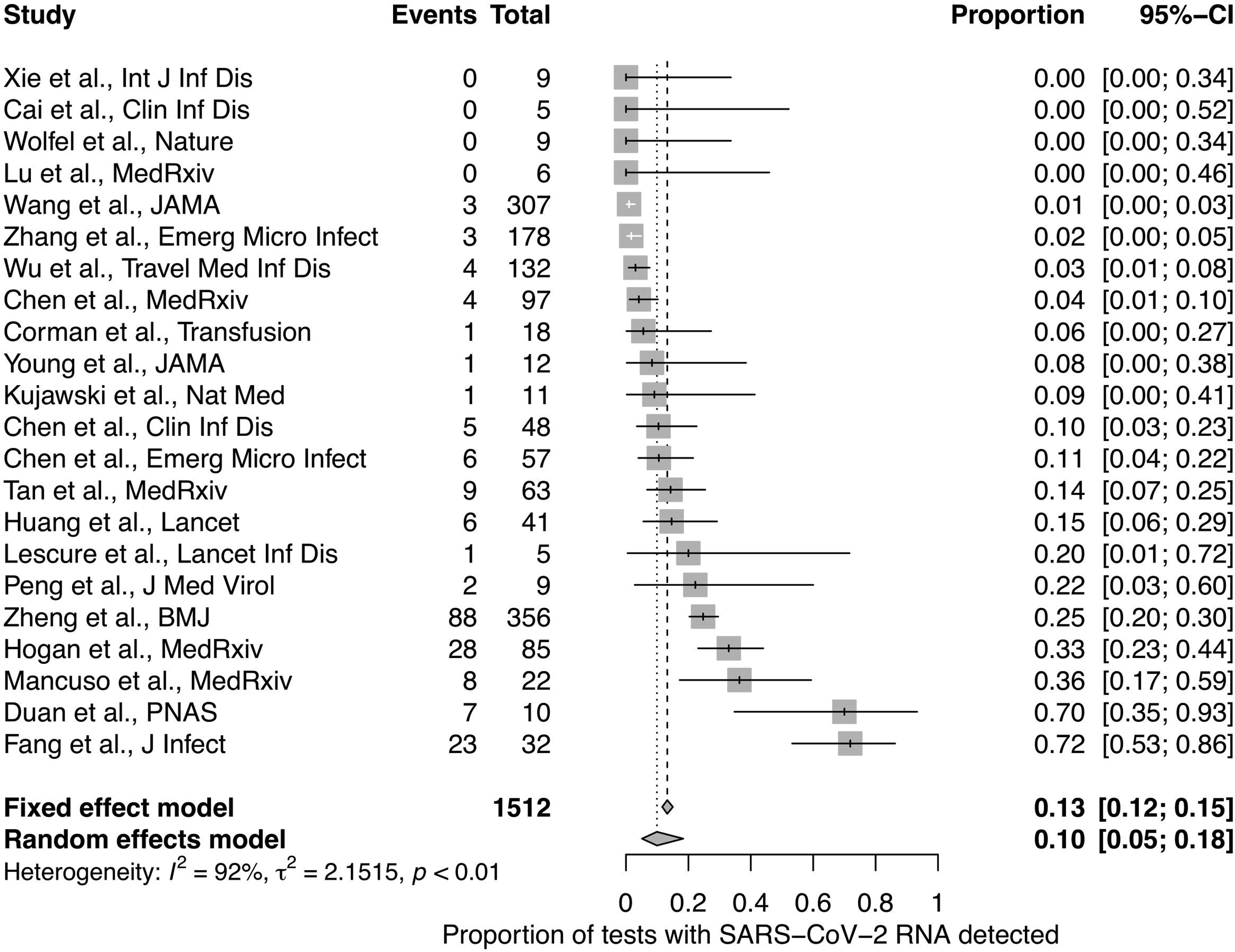
Prevalence of SARS-CoV-2 RNA in serum / plasma / whole blood samples from a systematic literature review. Point prevalence indicated for each study with confidence intervals showing citation and number of samples represented (Table 1).

### Cohorts and sample selection

We studied a total of 323 serum samples from 278 individuals (Table S2), all of whom had SARS-CoV-2 detected by a clinical diagnostic microbiology laboratory using RT-PCR on a respiratory swab. We collected 212 serum samples through the microbiology department at Oxford University Hospitals NHS Foundation Trust, OUH NHSFT, comprising adults with a diagnosis of COVID-19, classified in three groups as follows:

i. ***Hospital in-patients***, n=139 samples from 94 participants; these were collected from individuals admitted to OUH NHSFT, a tertiary referral centre in the South East of England, for treatment of COVID-19. Samples were collected between 1-5 days following admission to hospital or intensive care (whichever came later), a median of 8 days following symptom onset (range 1-37 days).
ii. ***Convalescent healthcare workers***, n=41 samples from 41 participants; these were collected from healthcare workers from OUH NHSFT, following a period of ≥ 7 days absence from work following a diagnosis of COVID-19, a median of 12 days following symptom onset (range 7-17 days).
iii. ***Convalescent patients***, n=32 samples from 32 participants; these were collected from patients presenting to OUH NHS FT followed up in the community, a median of 42 days following onset of COVID-19 symptoms (range 31-62 days).

Additional samples were collected through NHS Blood and Transfusion (NHSBT), as follows:

(vi) ***Convalescent plasma donors***, n=111 samples from 111 volunteer plasma donors, ≥28 days from recovery of symptoms.
(v) ***Healthy pre-pandemic controls***, n=5 samples from 5 independent healthy volunteer donors, collected prior to December 2019.

In groups (i)-(iii) >1 sample was obtained from 45 individuals, so our clinical dataset overall represents 167 unique individuals with COVID-19. Among these 167 individuals, we classified severity of illness as asymptomatic, mild, severe, or critical based on standard WHO criteria [9]. All serum samples were frozen in 0.5ml aliquots at −20°C.

### RT-PCR on serum samples

Following nucleic acid extraction, we used reverse transcription (RT)-PCR to amplify SARS-CoV targets from serum samples. Due to different pathways for patient recruitment and sample processing, PCR protocols varied by cohort (indicated in Table S2). Further specific details of each method are provided in supplementary material (S3) available on-line [10]. In brief:

1. Samples from acute hospital admissions and convalescent health care workers were processed by the OUH NHSFT clinical microbiology laboratory, using a Symphony Rotorgene protocol with an RNA-dependent RNA polymerase (RdRP) gene target, validated by Public Health England (PHE) for use on respiratory samples [11][12].
2. For convalescent OUH NHSFT patients, a nested PCR was undertaken using newly developed PCR primers [13].
3. Convalescent samples collected through NHSBT were analysed by Public Health England (Colindale), targeting either RdRp or a conserved region of the open reading frame (ORF1ab) gene of SARS CoV-2, together with detection of an assay internal control to monitor the extraction and RT-PCR processes. Reverse transcription and PCR amplification was performed on an Applied Biosystems 7500 FAST system.

High cycle threshold (Ct) values (>37.0) are often viewed as being non-specific in clinical diagnostic laboratories depending on the clinical situation. However, for research purposes we collected and reported all Ct values.

### Viral culture system

For viral culture, we used 20 serum samples, designated VC01-20 (identified in Table S2). VC01-16 comprised acute and convalescent samples that were RT-PCR positive, selected at random from our sample bank, representing samples from 12 individual patients (four individuals were represented at two timepoints), collected at 3-20 days following onset of symptoms. VC17-20 were pre-pandemic control samples. One further sample collected from a pre-pandemic NHSBT serum donation was used as media (VC21).

Samples VC01-20 were provided blinded for viral culture experiments. 50 μL aliquots of samples VC1-VC20 were separately added to 2.4 × 10^5^ Vero E6 cells (Cell Bank, Sir William Dunn School of Pathology, University of Oxford) in 24 well plates. Cells were propagated in Dulbecco’s Modified Eagle Medium (DMEM) supplemented with 10% foetal bovine serum (FBS). Virus growth assays were done in DMEM supplemented with 1% FBS, glutamine and penicillin/streptomycin, according to published methods [14]. In parallel, wells of the same number of cells were cultured in triplicate without virus challenge but with 50 μL control serum (VC21), or in duplicate with a stock of Victoria/01/2020 SARS-CoV-2 passage 4 (Oxford) at calculated ten-fold serial dilutions per well of 78, 7.8, 0.78 and 0.078 plaque forming units (pfu) in 50 μL of control serum (VC21).

Wells were observed daily for cytopathic effects (CPE), and 50 μL samples were taken for vRNA extraction on day 3 post-challenge. Where residual sample volumes permitted, 50 μL aliquots of the respective serum were processed in parallel. In addition, 1 × 10^8^ vRNA copies produced by *in vitro* transcription and quantified by droplet digital PCR were spiked into two equivalent control media samples, and processed in parallel, to provide quantification and estimate the loss of vRNA during extraction. All samples were processed for vRNA using QIAamp Viral RNA Mini kits according to the manufacturer’s instructions. RNA extracts were analysed by qRT-PCR, and vRNA copy number was interpolated from the standard curve of Ct value by known copy number. On day 4, 50 μL aliquots of supernatants from cells challenged with VC01-20 were “blind passaged” to fresh cells, and the remaining supernatants were harvested and stored separately at −80C for future analysis. After a further 3 days, we recorded CPE, if any, for second passage cultures.

### RT-PCR of culture supernatant

To determine whether there had been productive infection of cells *in vitro*, we took aliquots of culture supernatant, including positive and negative controls, and serum samples for qRT-PCR analysis using CDC NP1, CDC NP2 and HKU ORF1b diagnostic panels. RNA standards with a quantitative logarithmic range from 10^7^ to 10^1^ vRNA copies/reaction were included to calculate vRNA copy numbers across all samples. Details are provided in supplementary material S4.

### Ethics

Acute hospital in-patients were recruited into the Sepsis Immunomics study (Ref: 19/SC/0296). Convalescent healthcare workers with hospital encounters (n=38) and convalescent patients (n=32) provided informed consent for recruitment into the ISARIC WHO Clinical Characterisation Protocol UK (ISARIC WHO CCP-UK), with ethics approval by the South Central (Oxford C) Research Ethics Committee in England (Ref: 13/SC/0149), and Scotland A Research Ethics Committee in Scotland (Ref: 20/SS/0028). Additional convalescent healthcare workers were recruited by the Oxford GI Biobank, n=3 (approval by Yorkshire and The Humber - Sheffield Research Ethics Committee, ref. 16/YH/0247). Healthy pre-pandemic control samples were used under the NHSBT ethics, providing donor consent for their samples to be used in research.

### Statistical analysis

Anonymised data were stored using Microsoft Excel. We analysed and presented data using GraphPad Prism v.8.3.1. Statistical analyses were undertaken using R 3.6.2. Binomial confidence intervals are presented for proportions. Univariable and multivariable logistic regression models were used to determine associations between detectable vRNA and time since symptom onset, disease severity and patient sex and age, accounting for any non-linear effects of continuous factors using natural cubic splines. Meta-analysis was undertaken using the meta package for R, version 4.12.

## RESULTS

### Literature review to determine the frequency and clinical associations of RNA-aemia

We identified 28 relevant studies (Table 1; Table S1), among which 22 contained metadata suitable for meta-analysis (Fig 1). Point estimates for the frequency of vRNA detection are presented for each study representing ≥5 individuals, together with 95% confidence intervals (Figure 2; Table S1). We observed considerable heterogeneity in the range of estimates for vRNA-aemia, from 0% in several studies [15–19], up to 76% in a report of patients in a critical care setting [20]. Pooling the data from these reports, the point estimate for the prevalence of vRNA in blood products in the 28 days following symptomatic infection is 10% (95%CI 5-18%, random effects model).

**Table 1:** Frequency of SARS-CoV-2 RNA in human blood and blood products based on a systematic literature review. Full metadata are presented in S1 table, available online.

| Citation | Setting | Frequency and characteristics of SARS-CoV-2 RNA |
| --- | --- | --- |
| **ACUTE COVID-19 INFECTION** |  |  |
| Wang et al., *JAMA* [4] | n=205 patients with COVID-19; Hubei and Shandong provinces and Beijing, China | ☐ Blood: 3/307 samples RNA positive collected from 205 patients (0.98%); mean Ct 34-35<br>☐ No difference in Ct values between blood, stool, and respiratory samples |
| Zhang et al., *Emerging Microbes & Infections* [37] | n=178; Wuhan pulmonary hospital, China | • Whole blood: 6/178 (3.4%) PCR positive; Ct 30-32<br>• Serum: 3/178 (1.7%) PCR positive; Ct 24-33<br>• None of the patients with viral RNA detected in blood had positive respiratory swabs |
| Lescure et al., *Lancet Inf. Dis.* [21] | n= 5, hospital patients, France | • Plasma: 1/5 (20%) PCR positive; Ct >35<br>• Latest positive 12 days after symptom onset.<br>• The patient with vRNA-aemia was the most severely ill. |
| Duan et al., *PNAS* [38] | n= 10, severe COVID19 patients, Wuhan, China | • Serum: 7/10 (70%) PCR positive; Ct 34-38 |
| Chen et al., *CID* [22] | n=48, General Hospital of Central Theater Command, PLA, Wuhan, China | • Serum: 5/48 (10%) PCR positive<br>☐ RNAaemia only in the critically ill group (but 12 critically ill patients had no RNA-aemia)<br>• RNA-aemia associated with elevated IL-6 |
| Chen et al., *Emerg Microbes Infect.* [23] | n=57, Guangzhou Eighth People's Hospital, China | • Serum: 6/57 (11%) PCR positive; Ct 32-41<br>• RNA-aemia associated with severe symptoms |
| Fang et al., *J. Infect.* [20] | n=32, Central Hospital of Xiangtan, China | • Blood: 7/8 (88%) PCR positive in ICU patients and 16/24 (67%) in non-ICU patients. |
| Han et al., *CID* [39] | n=2, Seoul Metropolitan Government-Seoul National University, Korea | • Mother and 27 day old infant<br>• Plasma: RNA detected in infant up to day 10, mother's plasma negative |
| Huang et al., *Lancet* [40] | n=41, hospitalised patients, Jin Yin-tan Hospital, Wuhan, China | • Plasma: 6/41 (15%) PCR positive<br>• No difference in ICU admissions between patients with and without RNA-aemia. |
| Yu et al., *CID* [15] | n=4, Beijing Ditan Hospital, Capital Medical University, Beijing, China | • Blood: 0/4 (0%) PCR positive |
| Young et al.,<br>*JAMA* [41] | n= 18, hospitalized patients, Singapore | Blood: 1/12 (8%) PCR positive |
| Xie et al., *Int J Inf Dis.* [16] | n=9, Sichuan Provincial People's Hospital and Sichuan Mianyang 404 Hospital, Chengdu, China | Blood: 0/9 (0%) PCR positive |
| Wu et al.,<br>*Travel Med Inf Dis.* [42] | n=132, The East Section of Renmin Hospital of Wuhan University, China | Blood: 4/132 (3.03%) PCR positive |
| Cai et al., *CID* [17] | n=5, Childrens' hospital, Shanghai | Serum: 0/5 PCR positive within 2-3 days of symptom onset |
| Zheng et al. *BMJ* [3] | n= 96 admitted patients Zhejiang province, China | Serum: 39/96 (41%) overall (6/22 (27%) in mild cases, and 33/74 (45%) in severe case)<br>No difference in viral load between mild and severe cases<br>Serum had the lowest viral load compared with stool and respiratory samples. |
| Wolfel et al.,<br>*Nature* [18] | n=9, hospitalised, Munich, Germany | Serum: 0/9 (0%) PCR positive |
| Kujawski et al.,<br>*Nature Medicine* [24] | n=11, hospitalised patients, USA | Serum: 1/11 (9%) PCR positive<br>Detection of RNA in serum associated with clinical deterioration |
| Peng L et al., *J Med Virology* [43] | n=9, hospitalised patients, Sun Yat-sen University, China | Whole blood: 2/9 PCR positive |
| Corman VM et al., *Transfusion* [44] | n=18, range of patients, Germany | Serum: 1/18 PCR positive, in patient with ARDS needing mechanical ventilation.<br>SARS-CoV-2 present at 179 copies/ml |
| Song et al.,<br>*MedRxiv* [45] | n=1, China | Plasma: 0/1 positive |
| Lu et al.,<br>*MedRxiv* [46] | n=6, hospitalised patients, Jiangsu, China. | Serum: 0/6 positive |
| Mancuso et al<br>*MedRxiv* [47] | n=22 (10 severe disease, 12 mild disease), Milan, Italy | Plasma: 6/10 RNA positive in severe group (60%) and 2/12 (1.6%) in the mild group. |
| Hogan et al.,<br>*MedRxiv* [48] | n=85, California, USA | Plasma: 28/85 detectable RNA<br>Median Ct value 37.5 (compared with 27.1 for nasopharyngeal aspirate)<br>Those with RNA-aemia were older and more likely to go to ICU and need mechanical ventilation<br>All deaths occurred in those with RNA-aemia |
| Tan et al.,<br>*MedRxiv* [49] | n=67, Chongqing,<br>China | 9/63 (14%) positive for RNA |
| Chen et al.,<br>*MedRxiv* [50] | n=97, Zhuhai, China | Whole blood: 4/97<br>All 4 patients with RNA-aemia had the lowest oxygenation |
| Bouadma et al.,<br>*MedRxiv* [51] | n=1, Paris, France | Blood: 1/1 RNA detected<br>Patient developed multi-organ failure and died |
| **CONVALESCENT PATIENTS (>28 days)** |  |  |
| Ling et al<br>*Chinese Med J*<br>[19] | n=14, convalescent<br>patients | Serum: 0/14 (0%) |
| **HEALTHY DONORS** |  |  |
| Chang et al.,<br>*Emerging*<br>*Infectious*<br>*Diseases* [25] | n= 7425<br>Healthy blood<br>donors, Wuhan<br>Blood Center, China.<br>Collected Jan-March<br>2020, peak epidemic. | Prospective testing of 1,656 platelet donations and 774 whole blood donations: 1/2430 RNA positive (0.04%)<br>Retrospective testing of whole blood donations: 3/4995 RNA positive (0.1%) |

Viral RNA-aemia was reported in association with more severe disease in some studies, including a higher risk of admission to critical care settings, and increased incidence of acute clinical deterioration [21–24]. One study reported lower RNA levels in serum compared to other sample sites [3], whilst another found RNA levels in blood to be no different to that of other sample types [4]. In a small number of reports that included specific Ct values, these were typically high, although studies used variable PCR targets and different thresholds for reporting positivity (details of methods and reported Ct values are available in Table S2).

We excluded two studies from the meta-analysis because they focused on cohorts with different characteristics from all other samples sets. One of these reported PCR results from samples taken at timepoints beyond 28 days, among which none contained vRNA [19]. The other investigated vRNA-aemia in blood donors in Wuhan, China at the time of the peak of the local epidemic in the first three months of 2020, finding vRNA in six samples from among >7000 screened [25].

### Frequency and timing of SARS-CoV-2 RNA-aemia in a local cohort

Our local clinical sample set included n=212 samples from 167 patients (median age 57 years, IQR 46-76), 89 male (53%). In 163 patients in whom clinical data were available, disease was classified as asymptomatic (n=1, 0.6%), mild (n=81, 50.0%), severe (n=37, 22.7%), or critical (n=44, 27.0%). In this sample set, collected at a median of 11 days post symptom onset (IQR 7-17 days), 27/212 were PCR positive for vRNA (12.7%, 95%CI 8.6-18.0%). Deduplicating this to represent 167 unique individuals, 20 (12.0%, 95%CI 7.5-17.9%) had RT-PCR positive serum at any time point tested. Considering all 212 samples in a multivariable analysis, critical disease severity was associated with increased vRNA-aemia, comparing mild and asymptomatic cases to severe (OR 2.3, 95%CI 0.5-12.6, p=0.29) and critical cases (OR 7.5, 95%CI 2.0-37.3, p=0.006) (Table 2). Within this dataset there was moderate statistical evidence of a trend towards decreased odds of vRNA-aemia over time (OR, per day, 0.95, 95%CI 0.89-1.00, p=0.12) (Table 2; Figure 3).

**Table 2:** Odds ratios (OR) for associations between RNA-aemia and other patient characteristics, among 212 adults with confirmed COVID-19 infection recruited at Oxford University Hospitals NHS Foundation Trust.

| **Clinical attribute** | **Multivariable**<br>**OR** | **95% CI** |  | **P value** |
| --- | --- | --- | --- | --- |
| Age, per 10 years | 0.97 | 0.74 | 1.29 | 0.85 |
| Sex, Female | 1.00 | *(reference)* |  |  |
| Sex, Male | 1.54 | 0.61 | 4.10 | 0.37 |
| Severity, Mild or Asymptomatic | 1.00 | *(reference)* |  |  |
| Severity, Severe | 2.31 | 0.51 | 12.64 | 0.29 |
| Severity, Critical | 7.46 | 2.02 | 37.31 | 0.006 |
| Time from symptom onset, per day | 0.95 | 0.89 | 1.00 | 0.12 |

**Figure 3.**
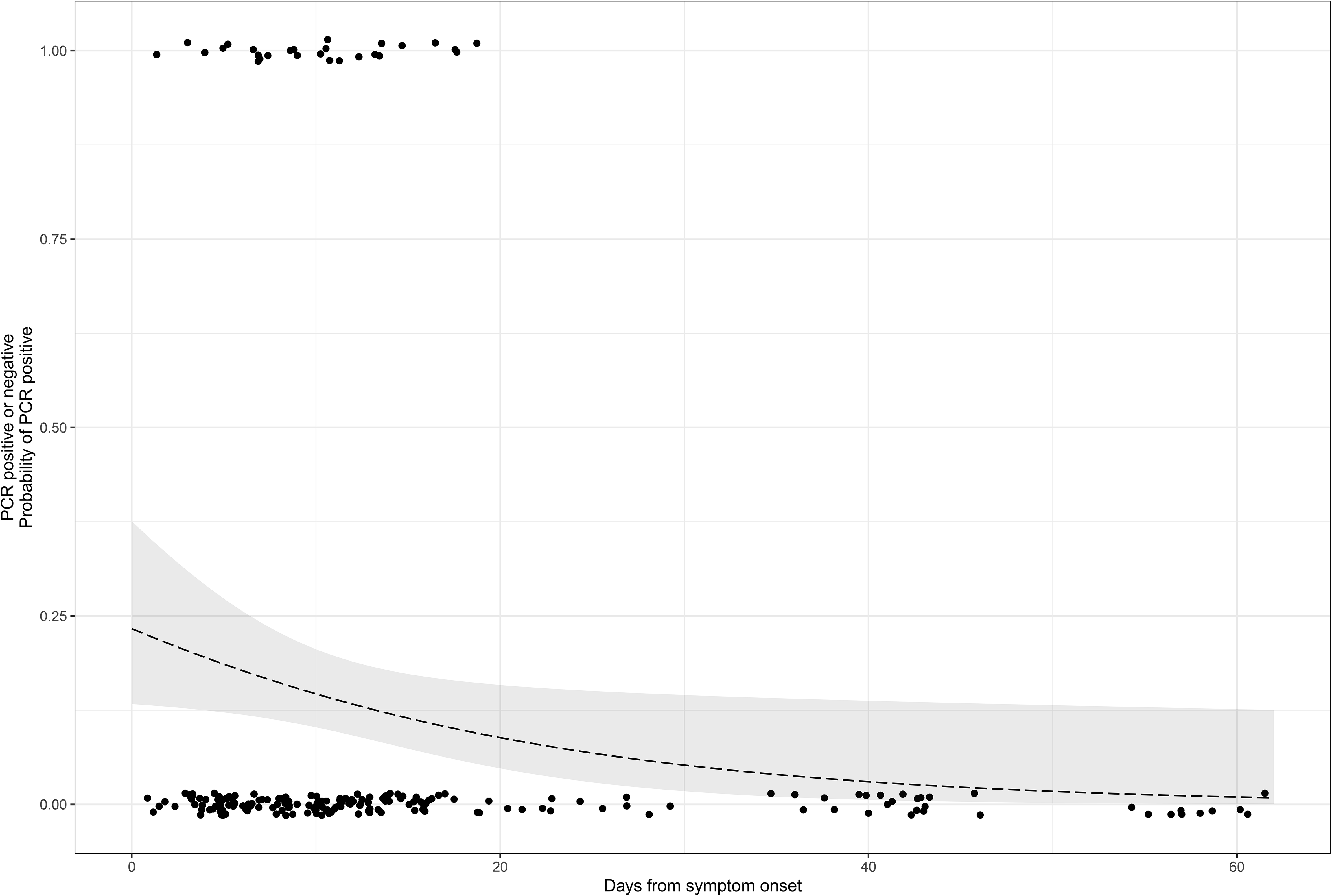
Relationship between RNA-aemia and days from COVID-19 symptom onset. Data shown for 212 samples collected from acute and convalescent adults from the Oxford University Hospitals cohort. Positive and negative results are shown plotted at 1 and 0 on the y-axis respectively, with jitter applied to show all points. The line shows the univariable predicted probability of RNA detection over time (95% CI: shaded).

Pooling our hospital data with results from the NHSBT convalescent cohort, vRNA was detected in 23/131 (17.6%, 95%CI 11.5-25.2) samples collected up to day 13, 4/40 samples from between day 14-27 (10.0%, 95%CI 2.8-23.7%), and 0/143 samples at ≥28 days (0%, 95%CI 0.0-2.5%) (Figure 2B). Day 20 was the latest time point at which any PCR positive sample was collected.

Ct values for all of our 27 PCR-positive sera were high (median 40.9, range 33.6-44.8). Using the more stringent Ct threshold of 37 that may be applied by clinical laboratories to report a positive result, only 7/25 fell below this cut off, reducing our overall positive rate to 7/212 among the local clinical cohort (3.3%, 95%CI 1.3-6.6%) or 7/424 across our entire sample set (1.6%, 95%CI 0.7-3.4%).

### Cytopathic effects arise in cell cultures inoculated with reference viral stock, but not in samples from COVID-19 patients or pre-pandemic controls

Healthy uninfected control cell cultures of Vero E6 cells were established (Figure 4A). We observed substantial cytopathic effects (CPE) in all samples inoculated with reference virus, characterised by cell rounding up and detaching (Figure 4B). CPE of this type was observed in wells challenged with 78 and 7.8 pfu, and moderate but typical CPE was observed in one well challenged with calculated 0.78 pfu reference virus. Cells exposed to a 1/10 dilution of control plasma did not show typical viral CPE. However in contrast to the CPE seen with reference virus, these control samples, the VC01-20 test cultures, and the culture inoculated with 0.078 pfu, instead showed variable cellular abnormalities and noticeable gel-formation in most samples (Figure 4C). Second passage cultures were undertaken in all cases, and none showed evident cytopathic effects at day 7 (Figure 4D). This approach is limited to the sample volume of 50 μL but we have demonstrated a single pfu at this volume reliably. Our detection threshold is <20 pfu/mL plasma, suggesting that plasma samples contain <0.1 infectious unit per 50 μL.

**Figure 4.**
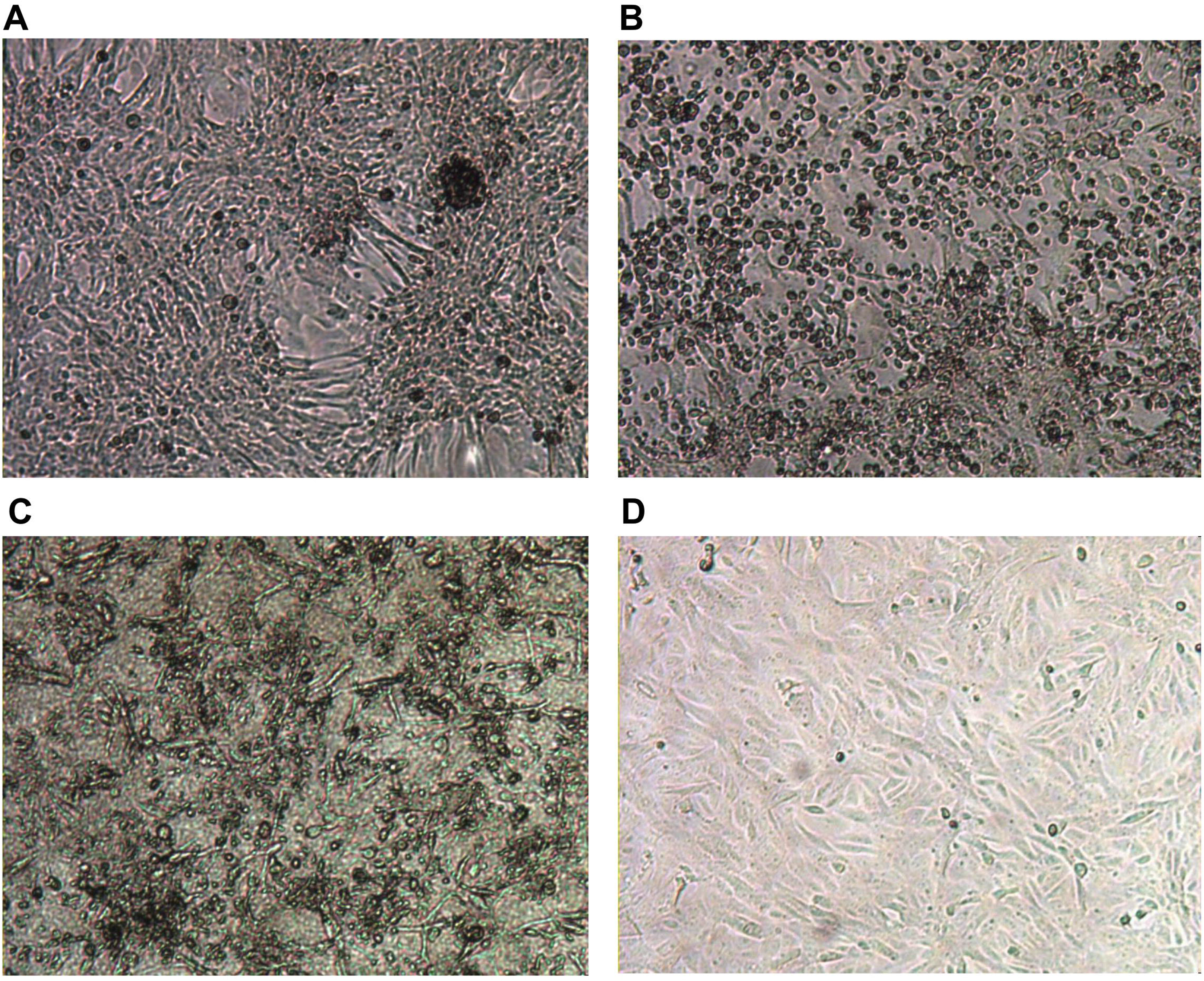
Typical images from cell culture in an *in vitro* system for SARS-CoV-2 culture. Top row shows controls: (A) Negative control Vero E6 cells in media; (B) Cytopathic effect (CPE) in Vero E6 cells spiked with Victoria/01/2020 SARS-CoV-2; Bottom row shows Vero E6 cells inoculated with 1/10 dilution of serum sample from sample VC12 (patient ID UKCOV040), that tested positive for SARS-CoV-2 RNA by RT-PCR; (C) Aberrant cellular effects at day 4 in a culture inoculated with VC12 at day 0; (D) Normal appearance of cells at day 7 inoculated with 1/10 dilution of the culture supernatant of the VC12-challenged culture, illustrated in (C).

### RT-PCR of culture supernatant

To determine whether there had been productive infection of cells *in vitro*, we took aliquots of culture supernatant for RT-PCR. From the positive control cultures, any culture receiving ≥1 infectious unit of virus (78 and 7.8 pfu in 50 μL of control serum) on day 0 produced ≥1 × 10^5^ copies of viral RNA (vRNA) per sample by day 3, detected by all three (CDC NP1, CDC NP2 and HKU ORF1b) primer/probe sets. All diagnostic panels also detected low levels of vRNA in the culture inoculated with a calculated dose of 0.078 pfu. These vRNA traces are likely to reflect fragments and RNA debris from the cells in which the virus was grown.

No serum sample, and no serum-inoculated cultures had >100 vRNA copies by day 3 based on the CDC, NP1 and NP2 assays. Marginal vRNA was detected in 10 serum samples, but none of these showed a rising titre by day 3 and none had vRNA levels within the reliably quantifiable range. The highest were in the range found in the sub-infectious dose positive control cultures (supplementary material S4). In contrast, no vRNA copies could be detected in serum-inoculated cultures tested at day 3 using the HKU ORF1b primer/probe set, while for the majority of original sera samples the HKU ORF1b assay gave similar results to the CDC, NP1 and NP2 diagnostic panel. The only exception from this was VC15, where marginal vRNA was detected in both serum and serum-inoculated cultures. These results suggest that no rising titre or no vRNA can be detected in serum-inoculated cultures. The comparison between the CDC and HKU diagnostic panels highlights interesting differences for detection of SARS-CoV-2 virus and should be explored further.

## DISCUSSION

### Impact of results

Recognition that SARS-CoV-2 RNA may be detected and quantified in blood highlights its potential provenance as a biomarker, but also raises concerns about safety for personnel handling samples in clinical and research environments. Protocols to underpin the safe handling of blood samples need to consider the best evidence for routes and risks of transmission in order to mandate safe laboratory practice, being informed by the nature of the samples and the specific task being undertaken (including any risk of aerosol generation), while also maintaining optimum cost effective workflow of clinical samples. Local risk assessments may currently result in disparate protocols being established by different organisations, but risk assessments should be proportionate, and - as far as possible - unified, and evidence-based. Developing new data to support laboratory practice is an important foundation for standardising practical guidelines.

Based on a systematic review of the literature, together with our own data, we estimate that SARS-CoV-2 RNA may be present at low copy numbers in ~10% of blood samples obtained from individuals with COVID-19 prior to day 28, most of which arise at earlier timepoints and in the setting of more severe disease. Despite being PCR-positive for vRNA, none of our clinical samples exceeded the threshold for viral infectivity.

### Relationship between viral load and disease phenotype

The Ct values reported in the literature and in our local samples are high, reflecting low copy numbers and suggesting that assays may be detecting genomic fragments rather than replication-competent virus in blood. However, it is also possible that intact virions are present, but that these are immune-complexed or otherwise neutralised, accounting for the lack of CPE in our culture system.

A previous study reported a decline in RNA-aemia in severe cases from 45% at the time of admission to 11% by week 4, and in mild cases from 27% to 0% over the same time period, although these differences did not reach statistical significance [3]. Viral load (measured by qRT-PCR) in respiratory samples has been correlated with disease severity [3,26,27]. Detection of vRNA in blood therefore may be more common in severe/critical disease as a result of higher viral loads overall, or specifically relating to a high burden of infection in the lungs leading to spill-over into the circulation, or reflecting the destruction of infected cells in the respiratory epithelium. Multisystem end-organ disease caused by SARS-CoV-2 could reflect systemic viral dissemination by blood or lymphatics (potentially with direct infection of lymphocytes), or may arise as a consequence of a sepsis syndrome triggered primarily by localised pulmonary infection [28]. Given the high Ct values for vRNA in blood, the identification of virus in the vascular compartment currently remains non-specific; further work is needed to understand its origins and significance, and to determine whether vRNA in the blood is innocuous or could contribute to immune dysfunction and the systemic inflammatory process.

Further work is needed to determine the bioburden and clinical significance of SARS-CoV-2 in other tissue types, for example in faeces [29,30]. Different clinical and laboratory infection control practices to be considered for specific sample types, to determine the frequency and duration of carriage and to assess whether infectious virus can be detected.

### Caveats and limitations

Datasets reported in the literature represent mostly a small number of carefully selected patients, typically in the acute hospital setting and therefore biased towards inclusion of more unwell patients meeting WHO criteria for severe or critical disease. Recognising that the field that is currently moving at pace, we elected to include papers from the pre-print server MedRxiv, for which peer review has not been undertaken. As a result, not all material included has undergone this quality assurance step. Published reports frequently do not include timing of sample collection relative to diagnostic respiratory samples and/or symptom onset, samples from individuals with trivial or absent symptoms are not well represented in existing studies and there are insufficient data to distinguish between frequency or quantification of vRNA present in whole blood, versus serum or plasma.

Due to the logistics of rapid recuitment of different patient groups through different pathways, RT-PCR methods varied by cohort, potentially introducing some variation in the sensitivity of detection. In our clinical samples, we adopted an inclusive approach to reporting detection of vRNA, by including samples with Ct values above those which would normally be called positive by a clinical diagnostic facility. This may lead to an over-estimation of the true prevalence of RNA-aemia in this sample group. Many previous publications do not report ct values and direct comparisons between datasets are therefore difficult.

The absence of CPE and amplification of vRNA must be considered within the constraints of the low sample volume (50 μL in each assay), and the limits of detection within the assays used. We tested serum samples after they had been subjected to a freeze/thaw cycle, which could also have potential influence on retrieval of infectious virus. However, as samples were frozen in accordance with standard laboratory operating protocols within a few hours of collection, we anticipate this would have a limited impact on viral replication capacity, as has been demonstrated previously for other viruses [31–33].

### Conclusions

Our data confirm that blood from COVID-19 patients may contain detectable RNA, but this arises in a minority of samples and is typically in low copy numbers, often outside the threshold that would be reported as positive in a clinical diagnostic laboratory. Based on evaluation of a small sample set, we have found no evidence to suggest that blood samples containing RNA could yield replication competent virus, suggesting a negligible risk of transmission of SARS-CoV-2 to healthcare workers and laboratory staff from handling such material. However, laboratory practice should be informed by guidance from Public Health England [34], CDC [35] and WHO [36]; individual risk assessment is important to account for the nature of the material being handled and the process being undertaken. Universal precautions and routine safety procedures should be carefully observed, not only to protect from COVID-19 infection but also to provide protection from other potential pathogens. Further data are needed to determine the extent to which serum PCR positivity for vRNA is useful as a diagnostic or prognostic marker in patients with COVID-19 infection.

## Data Availability

Supporting metadata are on-line at Figshare, DOI: 10.6084/m9.figshare.12278249

## SUPPLEMENTARY MATERIAL

Material in this section is available online at Figshare (DOI: 10.6084/m9.figshare.12278249) [10].

**S1 Table: Metadata table providing data for prevalence of SARS-CoV-2 RNA in blood and blood products based on a systematic literature review**. Details of 28 citations are shown, and the 22 studies included in quantitative meta-analysis are indicated.

**S2 Table: Metadata table providing underlying data for serum samples from adults with confirmed SARS-CoV-2 infection, based on RT-PCR nose/throat swab**. Sheet 1: samples obtained through patients recruited into a UK clinical cohort at Oxford University Hospitals NHS Foundation Trust (n=212). Cells highlighted in blue show follow-up samples collected from the same individual at different time points. Cells highlighted in orange show serum PCR positives. Sheet 2: samples obtained from convalescent donors a minimum of 28 days post resolution of symptoms, via NHS Blood and Transfusion, NHSBT (n=111).

**S3 Methods:** Details of RT-PCR reagents and cycling conditions for detection of SARS CoV-2 RNA.

**S4 Methods and data:** qRT-PCR quantification of vRNA from sera and viral culture assays. File contains methods for qRT-PCR and calculation of vRNA copy numbers, and qRT-PCR results in figure and table format.

**PRISMA checklist:** reporting for systematic review and meta-analysis

**STROBE checklist:** reporting for cohort studies

## ACKNOWLEDGEMENTS

This work uses data provided by patients and collected by the NHS. We are grateful to the frontline NHS clinical and research staff and volunteer medical students, who contributed to these data in challenging circumstances, and the generosity of the participants and their families for their individual contributions in difficult times. We thank the BRC Oxford GI Biobank which is funded by the National Institute for Health Research (NIHR) Oxford Biomedical Research Centre (BRC). Laboratory work for this study was also funded by the generous support of philanthropic donors to the University of Oxford’s COVID-19 Research Response Fund. We also acknowledge the IBD Cohort Investigators for support in the Oxford GI Biobank, and thank Marco Kaiser (GenExpress, Germany) for providing RNA standards and advice for qRT-PCR experiments. Thank you to Carla Wright and Rosie McMahon for administrative help.

## AUTHORSHIP CONTRIBUTIONS

**Data curation -** DWE, PCM, HH

**Resources -** CVA-C, JKB, MC, HH, SO, DJR

**Conceptualization -** DC, DWE, WJ, PCM, AJM, TP, NS

**Supervision -** MIA, EB, MC, DC, HH, KJ, TJ, PK, JK, PCM, GRS, MGS, TP, PS

**Ethics and governance -** DC, SJH, JK, PK, AJM, RJP, DJR, MGS, LT

**Methodology -** TB, TJ, WJ, PS, MZ,

**Laboratory work -** KA, TB, SB, KD, CD, LD, JGJ, SI, AJ, JM, MSO, JRa, JRu, SW

**Clinical recruitment -** JK, AJM, DS, MGS, LT

**Formal analysis -** TB, DWE, WJ, PCM

**Literature review -** LOD, SFL, ALM

**Writing - original draft preparation -** PCM

**Writing - review and editing -** DC, LOD, DWE, TJ, WJ, ALM, NS, HH, PS, MZ

**Project management -** SH, DC, PCM, NS

All authors read and approved the final manuscript

